# Classification of familial and non-familial ADHD using auto-encoding network and binary hypothesis testing

**DOI:** 10.1101/2025.08.15.25333792

**Authors:** Rahman Baboli, Elizabeth Martin, Qinyin Qiu, Lin Zhao, Tianming Liu, Xiaobo Li

## Abstract

Family history is one the most powerful risk factor for attention-deficit/hyperactivity disorder (ADHD), yet no study has tested whether multimodal Magnetic Resonance Imaging (MRI) combined with deep learning can separate familial ADHD (ADHD-F) and non-familial ADHD (ADHD-NF). T1-weighted and diffusion-weighted MRI data from 438 children (129 ADHD-F, 159 ADHD-NF, and 150 controls) were parcellated into 425 cortical and white-matter metrics. Our pipeline combined three feature-selection steps (t-test filtering, mutual-information ranking, and Lasso) with an auto-encoder and applied the binary-hypothesis strategy throughout; each held-out subject was assigned both possible labels in turn and evaluated under leave-one-out testing nested within five-fold cross-validation. Accuracy, sensitivity, specificity, and area under the curve (AUC) quantified performance. The model achieved accuracies/AUCs of 0.66 / 0.67 for ADHD-F vs controls, 0.67 / 0.70 for ADHD-NF vs controls, and 0.62 / 0.67 for ADHD-F vs ADHD-NF. In classification between ADHD-F and controls, the most informative metrics were the mean diffusivity (MD) of the right fornix, the MD of the left parahippocampal cingulum, and the cortical thickness of the right inferior parietal cortex. In classification between ADHD-NF and controls, the key contributors were the fractional anisotropy (FA) of the left inferior fronto-occipital fasciculus, the MD of the right fornix, and the cortical thickness of the right medial orbitofrontal cortex. In classification between ADHD-F and ADHD-NF, the highlighted features were the volume of the left cingulate cingulum tract, the volume of the right parietal segment of the superior longitudinal fasciculus, and the cortical thickness of the right fusiform cortex. Our binary hypothesis semi-supervised deep learning framework reliably separates familial and non-familial ADHD and shows that advanced semi-supervised deep learning techniques can deliver robust, generalizable neurobiological markers for neurodevelopmental disorders.

## 1. Introduction

Attention-deficit/hyperactivity disorder (ADHD) is among the most prevalent heterogeneous neurodevelopmental disorder (Faraone et al., 2021). Childhood ADHD, onsetting usually before the age of 12 years, affects approximately 5% of children worldwide (Polanczyk et al., 2014). Although many biological and environmental factors contribute towards risk of ADHD, family history emerges as the most robust risk determinant (Larsson et al., 2013; Samuel et al., 1999; Sprich et al., 2000). Children who have an affected first-degree relative (familial ADHD) face roughly a five-fold increase in risk compared with those without the history (non-familial ADHD) (Biederman et al., 1990). Moreover, familial cases are about four times more likely to exhibit persistent symptoms into adulthood (Biederman et al., 2012; Larsson et al., 2013; Samuel et al., 1999).

The neuroanatomical substrates distinguishing familial from non-familial ADHD remain poorly defined, and structural MRI studies have yet to establish a uniform anatomical profile for ADHD in general. Findings vary widely, with reports of volumetric reductions in fronto-striatal subcortical nuclei, putamen, caudate, thalamus, amygdala, and diminished cortical volume or surface area in frontal, parietal, and cerebellar regions (Batty et al., 2010; Hoogman et al., 2017; Mahone et al., 2011; Martine Hoogman et al., 2019), alongside large-scale analyses revealing widespread surface area deficits in frontal, cingulate, and temporal cortices (Martine Hoogman et al., 2019). Other studies describe focal cortical thinning in the pars opercularis, fusiform, parahippocampal, pre-central, and cingulate gyri, or delayed maturation of prefrontal and premotor cortices, whereas some report enlarged surface areas in medial superior-temporal and dorsomedial frontal regions or preserved limbic volumes (Batty et al., 2010; Yu et al., 2022). Investigation of long-range white matter pathways using diffusion tensor MRI (DTI) also reveals inconsistent results; many studies report lower fractional anisotropy (FA) or reduced tract volume in fronto-parietal, fronto-limbic, and other major tracts (Aoki et al., 2018; H.-L. Chiang et al., 2023; Hoogman et al., 2017; Peterson et al., 2011; Qiu et al., 2011), while others find higher FA or increased white matter volume in the same pathways (Chiang et al., 2016; Gau et al., 2015; Tung et al., 2021), and some large-sample studies detect no significant differences. Variation in acquisition and processing partly explains this heterogeneity, but the etiological diversity of ADHD is likely crucial. Familial risk, often unaccounted for, may contribute. Both children with ADHD and their unaffected first-degree relatives show reduced inferior, medial, and orbitofrontal volumes, suggesting heritable influences (He et al., 2015; Hoogman et al., 2017). However, without a non-familial ADHD comparison group, risk-specific neuroanatomy, whether in grey or white matter, remains unisolated. Overall, structural and microstructural studies confirm brain alterations in childhood ADHD, but the regions involved, and the direction of changes remain inconsistent, with familial risk potentially driving part of this variability.

Unlike traditional parametric methods, multivariate machine learning (ML) and deep learning algorithms can model the combined influence of large numbers of variables in high-dimensional space and are sensitive enough to uncover subtle patterns with strong discrimination or predictive value. Support-vector machines (SVMs) have primarily been used, and when combined with feature-selection or reduction methods such as recursive feature elimination, PCA, FFT, or ICA and validated with k-fold, hold-out, or leave-one-out cross-validation, SVM classifiers have separated children with ADHD from typically developing peers using structural MRI, resting-state functional Magnetic Resonance Imaging (fMRI) or DTI data (Cheng et al., 2012; Fair et al., 2013; Johnston et al., 2014; Sen et al., 2018; Yasumura et al., 2020). Salient discriminative features typically include functional connectivity of the thalamus and frontal, cingulate and temporal cortices, as well as cortical surface area, curvature, and voxel-based intensity measures (Brown et al., 2012; Colby et al., 2012; Iannaccone et al., 2015). Beyond SVMs, neural-network models (e.g., deep belief, convolutional and extreme learning networks) and other ML algorithms such as principal-component Fisher discriminant analysis, Gaussian-process classifiers and multiple-kernel learning have been trained on single- or dual-modality datasets with varying success (Aricò et al., 2020; Chaim-Avancini et al., 2017; Dey et al., 2012; Kumar et al., 2021; Loh et al., 2023; Qureshi et al., 2016; Qureshi et al., 2017; Zou et al., 2017). Despite these advances, most of these studies rely on a single imaging modality, omit systematic ranking of the most informative features, achieve only modest accuracies, and rarely examine systems-level structural or functional network properties or their links to clinical symptoms. Crucially, no published ML work has yet attempted to distinguish familial from non-familial ADHD, leaving the neurobiological signatures of these etiologically distinct subtypes unresolved.

In this study, we propose to utilize a deep learning technique, classification architecture based on binary hypothesis and using autoencoder, to identify the most robust structural and DTI brain signatures of children with and without familial risk for ADHD and controls. Deep learning frameworks excel at extracting latent feature sets by modeling the deep linear or nonlinear relationships throughout high dimensional measurement domains (LeCun et al., 2015). Based on findings from existing studies from our and other teams (Aoki et al., 2018; Baboli et al., 2023; Baboli et al., 2024; Baboli et al., 2025; Cao et al., 2023; Y. Luo et al., 2020; Luo et al., 2019; Saad et al., 2020; Xia et al., 2012) we hypothesized that structural topological alterations associated with frontal, parietal, and their interactions would significantly contribute to accurate discrimination of familial and non-familial ADHD and from controls.

## 2. Materials and methods

### 2.1. Participants

The present study analyzed de-identified baseline data from the Adolescent Brain Cognitive Development (ABCD) Study (Release 4.0), a nationwide cohort that enrolled 11 875 U.S. children aged 9–10 years across 21 sites through school-based, demographically stratified recruitment. The ABCD baseline resource integrates multimodal neuroimaging with an extensive suite of phenotypic data on both children and their parents including physical and mental health profiles, biospecimens, neurocognitive tests, substance-use histories, and environmental measures. Written parental consent and child assent were obtained under local IRB approvals, and the dataset is publicly available via the NIMH Data Archive. Children were eligible if they met all MRI-safety requirements, were fluent in English (parents could be fluent in English or Spanish) and scored ≥ 80 on the NIH-Toolbox Picture Vocabulary task. Children were excluded if they had a traumatic brain injury identified on the Modified Ohio State University TBI Screen– Short Version or any other neurological condition; a current or past diagnosis of bipolar disorder, schizophrenia, an autism-spectrum or pervasive developmental disorder, or chronic tic disorder; a chronic medical illness or use of non-stimulant psychotropic medication within the previous three months; or if any biological parent carried a diagnosis of autism, bipolar disorder, schizophrenia, or another psychotic disorder.

### 2.2. ADHD assessments

ADHD symptomatology was determined with the parent-reported, computerized Kiddie Schedule for Affective Disorders and Schizophrenia-5 (KSADS-5). Children who were marked according to the screening questions for a high risk of having an ADHD diagnosis were further administered with the KSADS-5 ADHD supplementary scales for diagnoses. The diagnosis of ADHD was based on the threshold of endorsing at least six out of nine symptoms of inattention (ADHD inattentive presentation), at least six out of nine symptoms of hyperactivity/impulsivity (ADHD impulsive/hyperactivity presentation), or those who met the criteria in both domains (ADHD combined presentation).

Familial loading was established from biological-parent Adult Self-Report (ASR) data; probands were placed in the familial subgroup (ADHD-F) if at least one parent carried a current/past ADHD diagnosis or scored > 65 on any DSM-5 ADHD-oriented ASR scale, and in the non-familial subgroup (ADHD-NF) when both parents were free of an ADHD diagnosis and scored < 60 on every ADHD-oriented ASR scale. Participants with insufficient information from both biological parents to determine positive vs. negative family history of ADHD were excluded. Typically developing controls endorsed no KSADS-5 ADHD symptoms and had parental ASR ADHD T-scores < 60. Sequential demographic, clinical and Human Connectome Project-based imaging quality filters yielded a final analytic sample of 438 children, 129 ADHD-F, 159 ADHD-NF and 150 controls. Control healthy subjects were matched to the ADHD-F and ADHD-NF cohorts on age, sex, handedness, estimated IQ, and pubertal category score.

### 2.3. Neuroimaging data acquisition protocol

MRI data were collected across the 21 ABCD sites on harmonized 3 T systems (Siemens Prisma, GE MR 750, or Philips Achieve), each fitted with a 32-channel receive array, following the consortium protocol detailed by Casey and colleagues (Casey et al., 2018). High resolution diffusion imaging was acquired with a single-shot spin-echo EPI sequence (TR/TE = 4100/88 ms on Siemens and 4100/81.9 ms on GE), 1.7 mm isotropic voxels, and 96 diffusion encoding directions distributed across four shells (b = 500 s mm⁻², 6 directions; b = 1000 s mm⁻², 15; b = 2000 s mm⁻², 15; b = 3000 s mm⁻², 60) acquired twice with reversed phase-encoding (AP/PA) over a 256 × 256 mm² field of view. Three dimensional T1-weighted volumes were obtained using an inversion prepared RF-spoiled gradient-echo sequence with 1.0 mm³ isotropic resolution, flip angle = 8°, and TR/TE pairs of 2500/2.88 ms (Siemens), 2500/2.0 ms (GE), and 6.31/2.9 ms (Philips); images were sampled on 176–225 sagittal slices within a 256 × 256 mm² (Siemens/GE) or 256 × 240 mm² (Philips) field of view. The present analyses used these diffusion and structural volumes after the ABCD consortium’s central preprocessing and quality control pipeline (Casey et al., 2018).

### 2.5 Individual-level image processing and feature generation

#### 2.5.1 Structural MRI

All T1-weighted images underwent a multi-stage visual quality assessment consistent with the Human Connectome Project structural QC protocol (Marcus et al., 2013). Each scan was assigned an overall score on a four-point scale, 1 (poor), 2 (fair), 3 (good) or 4 (excellent) based on sharpness, degree of motion blur and other artefacts. Datasets receiving a rating below 3 were excluded from subsequent analyses. The remaining data were processed in FreeSurfer v7.2.0 (Fischl, 2012; Zöllei et al., 2020), which briefly performs affine normalization to Talairach space, N4 bias-field correction, hybrid watershed skull stripping, tissue classification, and topology-corrected reconstruction of the white and pial cortical surfaces. Brain masks generated during skull stripping were inspected by two trained raters; minor defects such as residual dura was edited manually, whereas masks with major leakage led to exclusion of the subject. Following spherical registration to the fsaverage template, the cortex was parcellated into 34 gyral regions per hemisphere according to the Desikan-Killiany atlas, yielding regional thickness, surface area and curvature measures. Subcortical segmentation used a probabilistic atlas to label 37 nuclei after nonlinear alignment to the MNI305 template, and global morphometrics (intracranial volume, mean cortical thickness, total surface area and total subcortical volume) were also extracted. In total, 306 structural features, including regional cortical GM thickness, surface area, and GM volume of subcortical structures were extracted from each subject (**Figure 1**).

**Figure 1.**
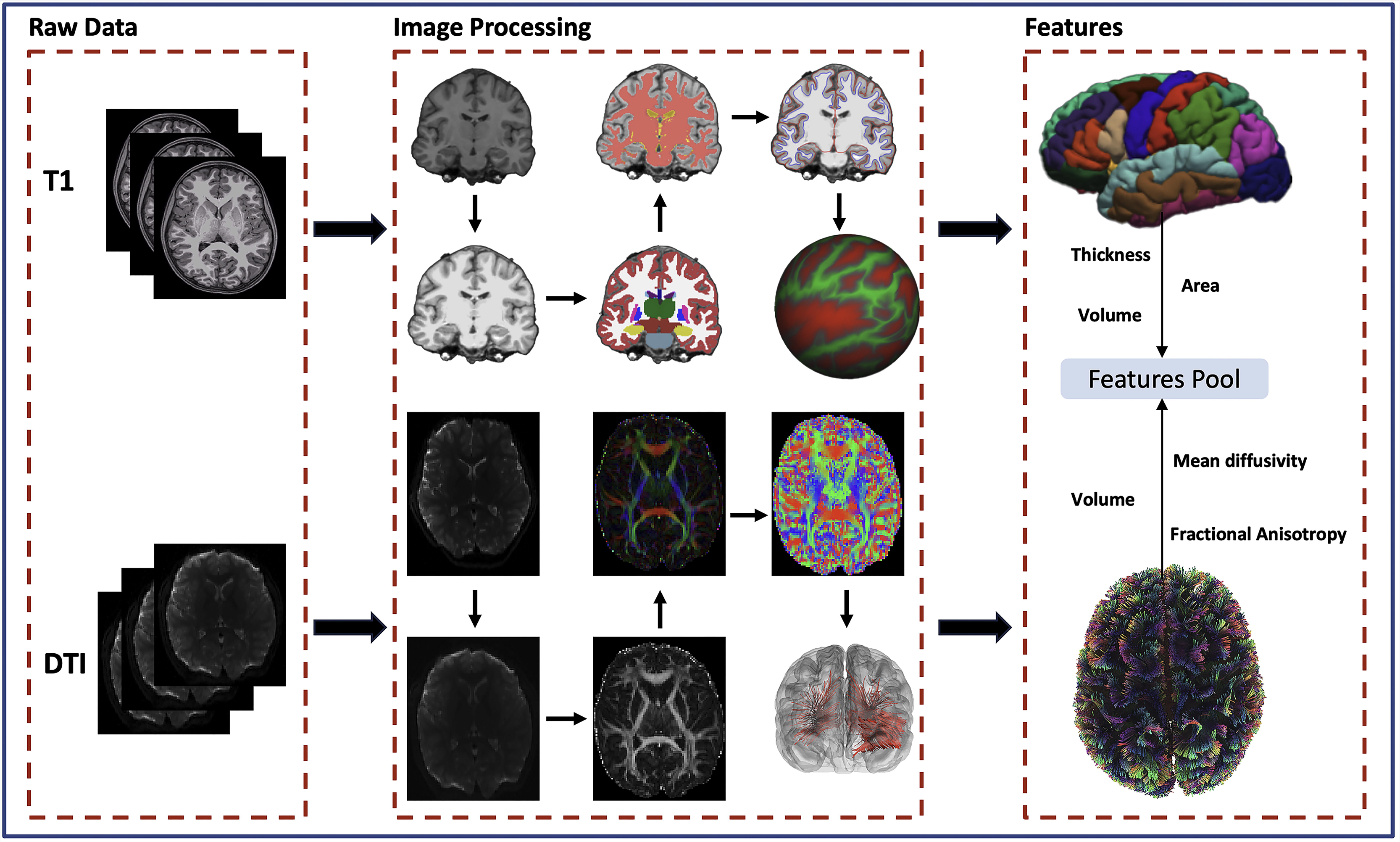
Multimodal preprocessing and feature-generation workflow.

#### 2.5.2 Diffusion-weighted Images

Diffusion-weighted images were examined volume-by-volume. Subjects with slice drop-out in more than two directions or head motion greater than 3 mm were removed. Pre-processing followed Human Connectome Project recommendations, beginning with susceptibility distortion correction using topup, then simultaneous eddy-current, motion correction and gradient reorientation with eddy, and finally rigid boundary-based registration of the corrected DWI to each participant’s T1 image to ensure anatomical congruence. Single-shell tensors (b = 1000 s mm⁻²) and multi-shell tensors (b = 1000/2000 s mm⁻²) were fitted with FSL’s dti-fit, generating fractional anisotropy, mean, radial and axial diffusivity maps. Whole brain tract segmentation employed AtlasTrack (Hagler Jr et al., 2009), which refines atlas fiber priors by comparing voxel-wise principal diffusion directions with subject specific tensors and labels 48 major association, projection, and limbic bundles. For each tract, mean FA, MD, RD, AD, and volume were extracted, and scan-level motion metrics were retained for quality control. The resulting diffusion feature set comprised 127 variables, microstructural indices for the 48 labelled tracts together with tract volumes and motion summaries. Each was standardized across participants before entry into the multimodal classification analysis designed to distinguish familial ADHD, non-familial ADHD and typically developing controls. In total, 128 DTI structural features, including the volume and FA of cortico-cortical and subcortico-cortical WM fiber tracts were extracted for each subject (**Figure.1**).

### 2.6 Binary-hypothesis classification framework

Our core classifier of our analyses follows the binary-hypothesis (BH) decision rule (Tang et al., 2021). For every outer fold iteration, the labelled training set is augmented twice for each held-out subject x: once under the provisional label b =1 (i.e., familial ADHD) and once under c =1 (i.e., non-familial ADHD). Within each augmented set the total within-class dispersion D_intra_ and the overall dispersion D_inter_ are computed [**Eq. (1)**, **Eq. (2)**].

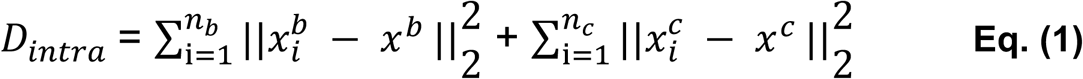

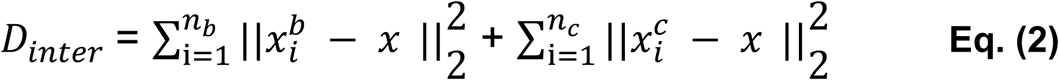

A variability ratio [**Eq. (3)**] then is obtained for both label assumptions, and the test subject is assigned to the class that minimizes this ratio.

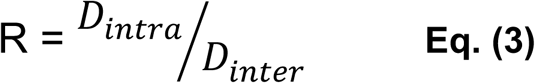

The procedure therefore prefers the assignment yielding the tightest within-class clustering relative to the global spread, exploiting distributional structure rather than proximity to a single centroid. To avoid circularity, feature ranking and, when selected the auto-encoder are re-trained inside a leave-one-out cross-validation loop that is nested entirely within the training data of each outer five-fold split.

#### 2.7.1 Feature selection

To limit over fitting while retaining the most informative neuroimaging markers, we combined three independent filter approaches, t-test filtering, mutual information (MI) ranking, and Lasso. MI ranking was used first because it captures non-linear statistical dependence between each variable and the class label, thereby favoring features that reduce label uncertainty beyond what linear metrics can reveal (Vergara & Estévez, 2014). A Lasso feature selection method was trained; the ℓ₁ regularize forces many weights to zero, producing a sparse subset that still maximizes the separating margin and has proved effective in high-dimensional biomarker discovery (Moon & Nakai, 2016). The top twenty scoring variables from each method were pooled, yielding at most sixty candidate features, out of the original 425 gray- and white-matter measurements. For every training split, all retained features were z-standardised (mean 0, SD 1) before they entered the auto-encoder embedding and subsequent binary-hypothesis classification.

#### 2.7.2 Modeling of autoencoder

Standard auto-encoder (AE) was implemented in the TensorFlow deep learning framework via the Keras application-programming interface. The network ingests a z-standardised vector of union features selected inside each training split and passes it through an encoder comprising a fully connected hidden layer with a rectified linear unit (ReLU) activation (40 units) followed by a 20-node latent layer, also ReLU activated (Nair & Hinton, 2010) . A mirror symmetric decoder then produces a linear reconstruction of the input. All weights were initialized with the Glorot uniform scheme and optimized with the adaptive-moment-estimation (Adam) optimizer at a learning rate of 1 X 10^-3^ (Kingma & Ba, 2014). The training objective was the mean squared error (MSE) reconstruction loss minimized over 30 epochs with 10% of the training data held out for validation [**Eq. (4)**].

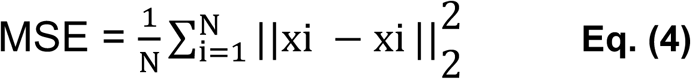

To avoid over- or under compression, a lightweight hyper parameter search tested three latent/hidden configurations (10/20, 20/20, 20/40). The setting that produced the lowest validation loss was retained. The resulting encoder generated fold-specific latent codes that supplanted the raw inputs in downstream binary hypothesis classification. Because the AE was re-trained inside every leave-one-out loop nested within the outer five-fold cross validation, no information leaked from the evaluation partition into feature learning.

### 2.8. Evaluation measures

To provide a multidimensional view of classifier performance, we used classification accuracy, sensitivity, specificity as well as AUC [**Eq (5)**, **Eq (6)**, **Eq (7)**, **Eq (8)**].

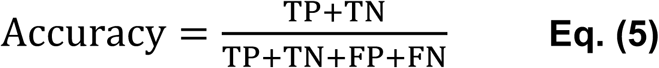

Because accuracy can mask class-imbalance effects, we also reported class-conditional metrics. Sensitivity (recall) gauges the model’s ability to recover true positives,

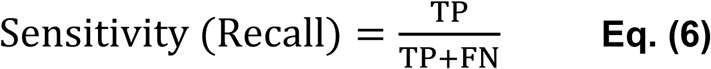

while specificity quantifies how effectively true negatives are recognized.

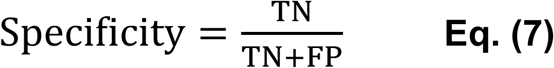

In addition, discrimination across all decision thresholds was summarized with AUC.

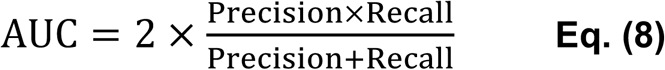

Figure. 2 presents the flow diagram of our analysis pipeline within a single fold of the five-fold cross-validation. For each held-out test subject, the entire pipeline runs twice. In the first pass, the subject is provisionally assigned to Group 1 (e.g., ADHD-F), undergoes feature selection, and passes through the auto-encoder, yielding a R1. In the second pass, the same subject is provisionally assigned to Group 2 (e.g., ADHD-NF), incorporated into the Group 2 training set, and processed through the identical feature-selection and auto-encoder steps to generate R2. The predicted label is the group that produces the smaller R. This two-pass test is repeated for every subject in a leave-one-out loop nested within the five-fold cross-validation. Accuracy, sensitivity, specificity, and AUC are then averaged across the five outer folds to produce the final performance metrics.

**Figure 2.**
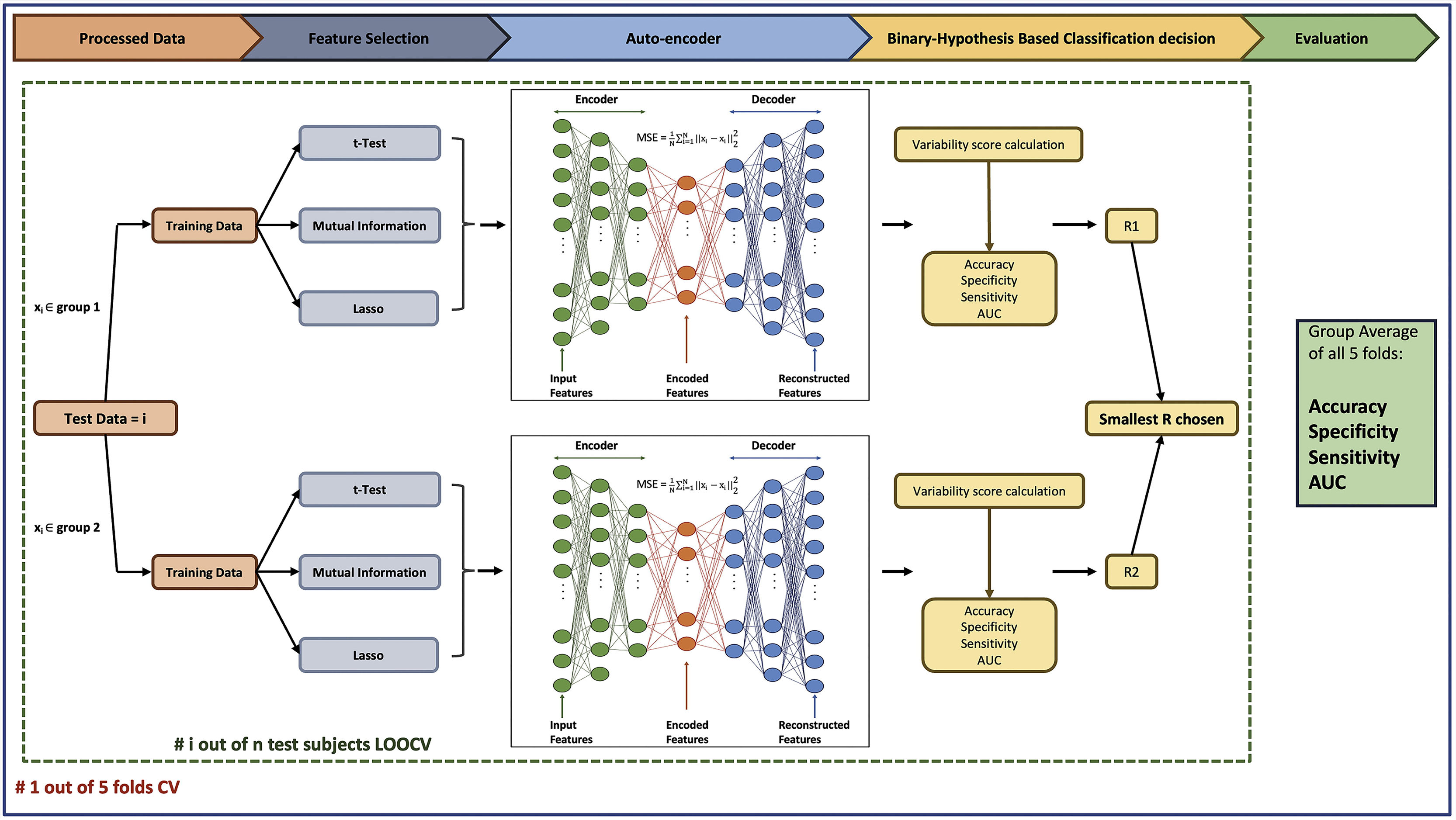
Pair-wise classification frameworks: For every held-out test subject (xi), the full pipeline is executed twice. In the first pass, the subject is assumed to belong to Group 1 (e.g., ADHD-F) and is taken through feature selection and an auto-encoder to produce R₁. In the second pass, the same subject is assumed to belong to Group 2 (e.g., ADHD-NF), is included with the Group 2 training pool, and the identical feature-selection and auto-encoder sequence is run to produce R₂. The smaller R designates the predicted label. This two-pass evaluation is applied sequentially to every test subject within a leave-one-out loop nested in five-fold cross-validation; accuracy, sensitivity, specificity, and AUC are averaged across the five outer folds to obtain the final performance metrics.

### 2.9. Feature importance score calculation

To quantify the contribution of each brain feature to the classification performance, we employed a permutation-based importance methodology (Breiman, 2001). This model-agnostic technique assesses a feature’s significance by measuring the degradation in model performance after its values are randomly permuted across all samples. A larger increase in model error implies a greater importance for that feature. For our classification model, the change in performance was specifically evaluated using the binary cross-entropy loss (Fisher et al., 2019; Strobl et al., 2008). The feature importance score for each feature was calculated using the following equation [**Eq. (9)**]:

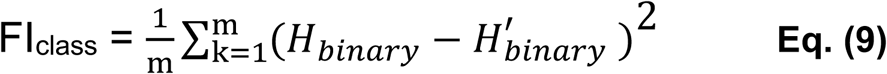

In this equation, m is the number of shuffling iterations, *H_binary_* represents the binary cross-entropy of the model on the original, unalerted dataset, and *H’_binary_* is the cross-entropy after a single feature’s values have been shuffled. In our analysis, we set m to 1,000 permutations to ensure a stable estimate. Following the calculation, a feature was identified as significant if its importance score was more than two standard deviations above the mean importance score of all input features (Guyon & Elisseeff, 2003; Sun et al., 2020).

## 3. Results

The demographic and clinical characteristics of the study participants are displayed in Table 1. The groups differences did not reveal any significant between-group differences among the ADHD-F, ADHD-NF, and control groups in terms of age, sex, handedness, puberty category score, IQ, race, income, parental education, and ADHD presentations. An analysis of medication usage revealed a similar distribution between the ADHD-F and ADHD-NF subgroups (p=0.715). The majority of ADHD (-F and -NF) participants were currently not on medication. Those who were on medication were primarily on stimulant medications, and very few individuals were receiving non-stimulant or combination medication therapies (see Table 1).

**Table 1.**
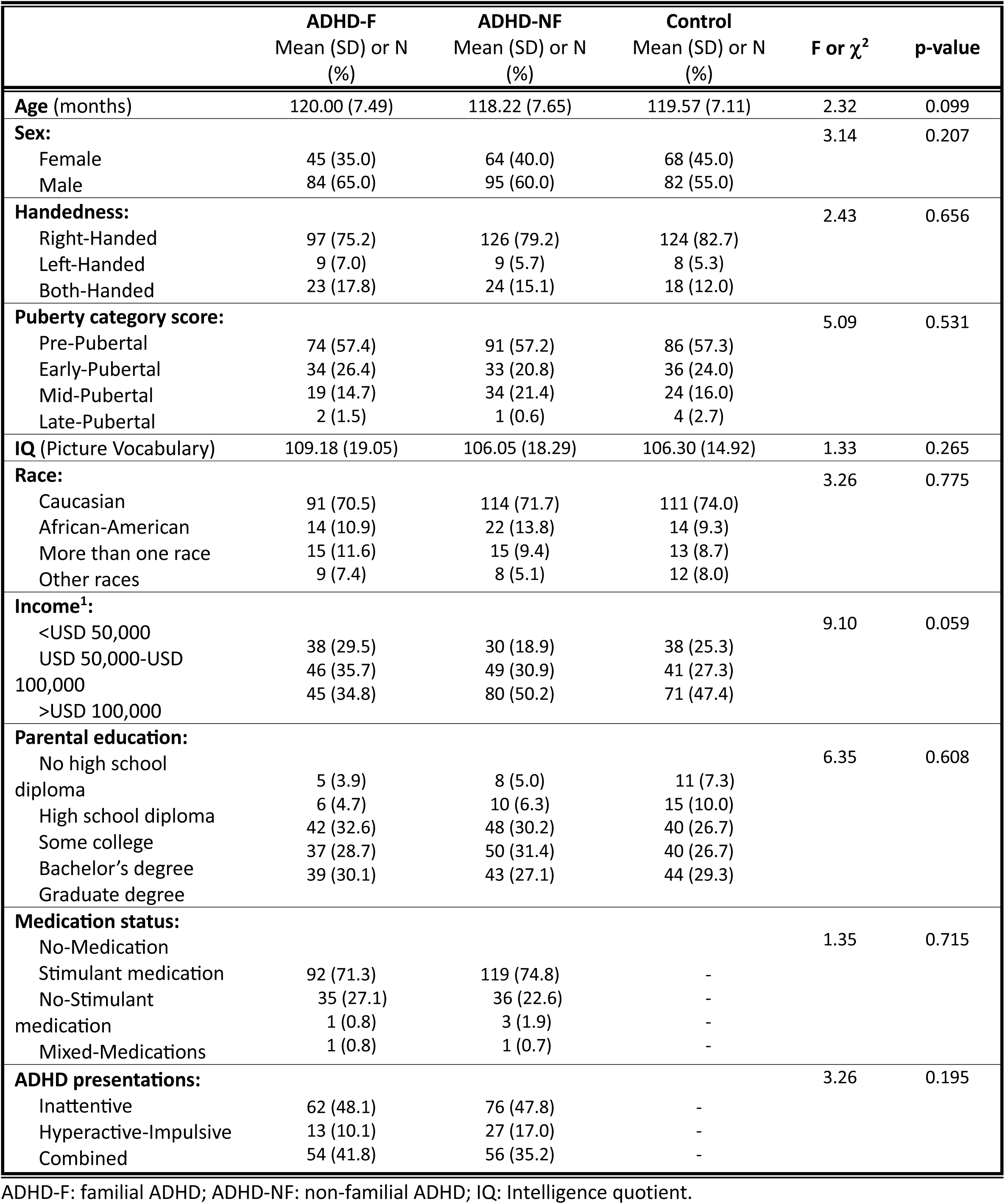
Demographic and clinical characteristics of the study cohort.

The classification performance of the Binary Hypothesis framework for the three group comparisons is summarized in Table 2. In discriminating between the ADHD-F and ADHD-NF subgroups, the model yielded an accuracy of 0.62, a sensitivity of 0.67, and an Area Under the Curve (AUC) of 0.67. For the classification between the ADHD-F group and controls, the model demonstrated a higher accuracy of 0.66, a sensitivity of 0.81, and an AUC of 0.67. The framework achieved its best overall performance when distinguishing ADHD-NF participants from controls, resulting in the highest accuracy (0.67), a sensitivity of 0.70, and the highest AUC (0.70).

**Table 2.** The results of classification performances between ADHD-F vs. ADHD-NF, ADHD-F vs. controls, and ADHD-NF vs. controls.

| Classification Performances | ADHD-F vs. ADHD-NF | ADHD-F vs. Control | ADHD-NF vs. Control |
| --- | --- | --- | --- |
| **Specificity** | 0.58 | 0.60 | 0.63 |
| **Sensitivity** | 0.67 | 0.81 | 0.70 |
| **Accuracy** | 0.62 | 0.66 | 0.67 |
| **AUC** | 0.67 | 0.67 | 0.70 |

The most predictive features for the classification of controls versus the ADHD-F group were mean diffusivity of right fornix, mean diffusivity of left para hippocampal cingulum, and thickness of right inferior parietal cortex. For the classification of controls versus the ADHD-NF group, the three most important characteristics were fractional anisotropy of left inferior-fronto-occipital fasiculus, mean diffusivity of right fornix, and thickness of right medial orbitofrontal cortex. Finally, discriminating between the ADHD-F and ADHD-NF subgroups was most influenced by volume of left cingulate cingulum tract, volume of right parietal superior longitudinal fasiculus tract, and thickness of right fusiform. The importance scores for these top-ranking features across all three classifications are presented in detail in Table 3.

**Table 3.**
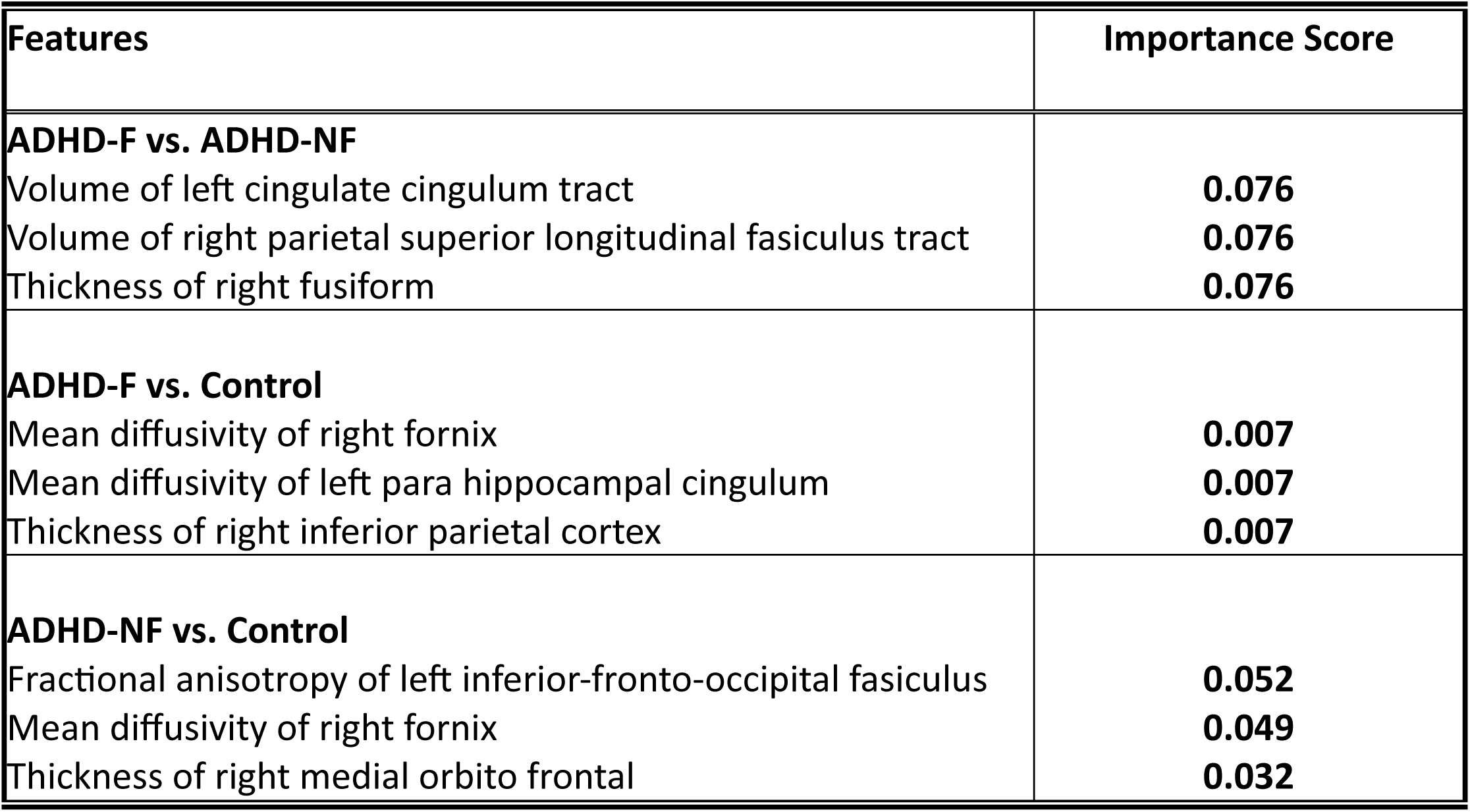
Importance scores of top six important features in classification distinguishing children between ADHD-F and ADHD-NF, ADHD-F and controls as well as ADHD-NF and controls.

## 4. Discussion

To our best knowledge, this is the first study to deploy a deep-learning framework on multimodal structural and diffusion neuroimaging data to identify the neural signatures that distinguish familial from non-familial ADHD in children, as well as ADHD-F versus controls and ADHD-NF versus controls. By training an auto-encoder under a binary-hypothesis approach on 425 cortical and white-matter metrics from 438 children (129 ADHD-F, 159 ADHD-NF, 150 controls), we isolated three most predictive brain features for each comparison. In the ADHD-F versus control analysis, the key variables were mean diffusivity of the right fornix, mean diffusivity of the left parahippocampal cingulum, and cortical thickness of the right inferior parietal lobule. For ADHD-NF versus controls, fractional anisotropy of the left inferior fronto-occipital fasciculus, mean diffusivity of the right fornix, and cortical thickness of the right medial orbitofrontal cortex provided the strongest discrimination. Finally, separating ADHD-F from ADHD-NF was chiefly driven by volume of the left cingulate cingulum, volume of the right parietal segment of the superior longitudinal fasciculus, and cortical thickness of the right fusiform gyrus. A subsequent binary-hypothesis classifier achieved accuracies between 0.62 and 0.67 across all pairwise contrasts, confirming that these complementary feature sets are sufficient to distinguish the subtypes and controls.

Our current analysis highlighted mean diffusivity in the right fornix and left parahippocampal cingulum, together with cortical thickness of the right inferior parietal lobule (IPL), as the strongest features for distinguishing the familial ADHD subgroup from typically developing controls. Limbic pathways such as the fornix and parahippocampal cingulum are integral to hippocampal–prefrontal communication; diffusion abnormalities in these tracts have previously been linked to episodic-memory deficits and increased inattentive symptoms in pediatric ADHD cohorts (Chiang et al., 2016; H. L. Chiang et al., 2023; González-Madruga et al., 2022). A large diffusion study of familial versus non-familial ADHD, likewise reported elevated diffusivity in the fornix of children carrying a heritable risk, suggesting that altered limbic microstructure may be a stable endophenotype rather than a state marker (Baboli et al., 2025). The IPL anchors the dorsal attention network, and convergent functional-MRI and machine-learning work shows that alterations in parietal hubs can reliably separate ADHD and control groups) (Brown et al., 2012; Colby et al., 2012; Fair et al., 2013; Fair et al., 2010). Our model confirms that abnormalities in the right IPL, in concert with limbic diffusivity metrics, are the most informative neural predictors for classifying familial ADHD.

Our binary-hypothesis pipeline revealed that fractional anisotropy of the left inferior fronto-occipital fasciculus (IFOF), mean diffusivity of the right fornix, and thickness of right medial orbitofrontal cortex (mOFC) were the most predictive variables for discriminating non-familial ADHD from typically developing controls. The IFOF is a major ventral stream tract that integrates visual salience with fronto-striatal control, and reduced FA in this pathway has been linked to heightened distractibility and slower attentional in children with ADHD (Konrad et al., 2006; van ’t Ent et al., 2009). In line with our finding that abnormalities in the medial orbitofrontal cortex occur together with reduced integrity of the IFOF, previous machine learning analyses reported likewise rank of ventral-frontal connectivity, including OFC and IFOF. These results support the view that environmental factors, rather than genetics, can impair the ventral attention network. Longitudinal studies suggest that ventral-stream measures normalize in children whose ADHD remits provide further evidence (Yuyang Luo, Tara L. Alvarez, et al., 2020; Yuyang Luo, Jeffrey M. Halperin, et al., 2020). In contrast to the importance of fornix diffusivity in both familial and non-familial comparisons, which suggests that limbic dysregulation is a shared substrate of ADHD, the selective involvement of the IFOF and mOFC in the non-familial group points to a network that may respond to environment-based treatment strategies such as reducing classroom visual noise or incorporating mindfulness training.

The familial versus non-familial contrast in our binary hypothesis model was predominantly driven by features located in the left cingulate cingulum and the right parietal segment of the superior longitudinal fasciculus for ADHD-F, alongside features in the right fusiform gyrus for ADHD-NF. These findings align with prior familial risk imaging work from our group, which has repeatedly linked cingulate cortex and SLF morphometry to hereditary liability and to the elevated persistence of symptoms in ADHD-F(Baboli et al., 2023; Baboli et al., 2025). The cingulate cingulum is a core conduit of the limbic– executive network. Greater tract volume in ADHD-F aligns with twin and sibling data showing heritable disruptions in anterior cingulate morphology and its fronto-striatal connectivity (Castellanos et al., 2003; Chen et al., 2018; Cheng et al., 2012). Meanwhile, the SLF-P links dorsal parietal attention hubs with the prefrontal cortex; its enlargement in familial cases echoes reports of heightened SLF centrality in children who remain symptomatic into adolescence and supports the hypothesis that genetic risk fosters a more homogeneous “executive-network” phenotype. By contrast, fusiform thinning in ADHD-NF is consistent with the environmentally modulated heterogeneity posited by large ENIGMA-ADHD meta-analyses, which associate fusiform and occipito-temporal anomalies with variable reading exposure and screen use rather than with family history (Yuyang Luo, Tara L. Alvarez, et al., 2020; Martine Hoogman et al., 2019). The divergence between limbic–parietal involvement in ADHD-F and ventral-stream involvement in ADHD-NF therefore reinforces recent evidence that heredity narrows anatomical variance, whereas non-familial ADHD exhibits broader, context-sensitive profiles (Mu et al., 2023). Collectively, these results strengthen the argument that familial ADHD represents a biologically more homogeneous subgroup anchored in limbic– executive circuitry, while non-familial ADHD remains anatomo-functionally diverse, an observation with direct implications for tailoring interventions and for stratifying future imaging cohorts.

The findings of this study should be interpreted in light of several limitations. First, although our cohort of 438 children is comparatively large for pediatric neuroimaging research, it remains modest for deep learning applications; even with mutual-information filtering, Lasso regularization, and five-fold cross-validation, some risk of over fitting and limited generalizability persists. Second, only one biological parent or guardian was interviewed and assessed during the study visits. Therefore, we excluded the children with ADHD who had insufficient information from both biological parents to determine positive vs. negative family history of ADHD in the groups of ADHD-F and ADHDNF. Second, all data were drawn from the baseline wave of the ABCD community sample, and ADHD diagnoses were based on parent-reported KSADS interviews rather than on clinician confirmed assessments. Therefore, findings of this study need to be validated in a clinical sample.

In conclusion, this study is the first to apply a binary hypothesis framework within a deep learning approach to multimodal structural and diffusion MRI data for identifying the most informative neural features that distinguish between children with familial ADHD, non-familial ADHD, and matched controls. By classifying each pair of groups, limbic–executive network structures, including the fornix, parahippocampal cingulum, cingulate cingulum, and superior longitudinal fasciculus, consistently emerged as the strongest predictors separating familial ADHD from the other groups, suggesting a heritable neural signature linked to symptom persistence. In contrast, ventral stream pathways such as the inferior fronto-occipital fasciculus, medial orbitofrontal cortex, and fusiform gyrus were key discriminators for non-familial ADHD, pointing to greater influence of environmental factors and a more heterogeneous neurobiological profile. By showing that distinct sets of structural and microstructural features can reliably classify each group contrast, our findings highlight the value of incorporating familial risk into neuroimaging research for improving biomarker precision and informing targeted intervention strategies.

## Data Availability

All data produced in the present study are available upon reasonable request to the authors

https://abcdstudy.org/about/

## Acknowledgment

This work was partially supported by research grants from the National Institute of Mental Health (R15MH117368) and the New Jersey State Department of Health (CBIR17PIL012, CBIR22PIL022). Data used in the preparation of this article were obtained from the Adolescent Brain Cognitive Development^SM^ (ABCD) Study (https://abcdstudy.org), held in the NIMH Data Archive (NDA). This is a multisite, longitudinal study designed to recruit more than 10,000 children age 9-10 and follow them over 10 years into early adulthood. The ABCD Study® is supported by the National Institutes of Health and additional federal partners under award numbers U01DA041048, U01DA050989, U01DA051016, U01DA041022, U01DA051018, U01DA051037, U01DA050987, U01DA041174, U01DA041106, U01DA041117, U01DA041028, U01DA041134, U01DA050988, U01DA051039, U01DA041156, U01DA041025, U01DA041120, U01DA051038, U01DA041148, U01DA041093, U01DA041089, U24DA041123, U24DA041147. A full list of supporters is available at https://abcdstudy.org/federal-partners.html. A listing of participating sites and a complete listing of the study investigators can be found at https://abcdstudy.org/consortium_members/. ABCD consortium investigators designed and implemented the study and/or provided data but did not necessarily participate in the analysis or writing of this report. This manuscript reflects the views of the authors and may not reflect the opinions or views of the NIH or ABCD consortium investigators.

## References

Aoki, Y., Cortese, S., & Castellanos, F. X. (2018). Research Review: Diffusion tensor imaging studies of attention-deficit/hyperactivity disorder: meta-analyses and reflections on head motion. Journal of Child Psychology and Psychiatry, 59(3), 193–202. 10.1111/jcpp.12778

Aricò, M., Arigliani, E., Giannotti, F., & Romani, M. (2020). ADHD and ADHD-related neural networks in benign epilepsy with centrotemporal spikes: A systematic review. Epilepsy Behav, 112, 107448. 10.1016/j.yebeh.2020.107448

Baboli, R., Cao, M., Halperin, J. M., & Li, X. (2023). Distinct Thalamic and Frontal Neuroanatomical Substrates in Children with Familial vs. Non-Familial Attention-Deficit/Hyperactivity Disorder (ADHD). Brain Sciences, 13(1), 46. https://www.mdpi.com/2076-3425/13/1/46

Baboli, R., Cao, M., Martin, E., Halperin, J. M., Wu, K., & Li, X. (2024). Distinct structural brain network properties in children with familial versus non-familial attention-deficit/hyperactivity disorder (ADHD). Cortex, 179, 1–13. 10.1016/j.cortex.2024.06.019

Baboli, R., Wu, K., Halperin, J. M., & Li, X. (2025). White Matter Microstructural Abnormalities in Children with Familial vs. Non-Familial Attention-Deficit/Hyperactivity Disorder (ADHD). Biomedicines, 13(3), 676. https://www.mdpi.com/2227-9059/13/3/676

Batty, M. J., Liddle, E. B., Pitiot, A., Toro, R., Groom, M. J., Scerif, G., Liotti, M., Liddle, P. F., Paus, T., & Hollis, C. (2010). Cortical Gray Matter in Attention-Deficit/Hyperactivity Disorder: A Structural Magnetic Resonance Imaging Study. Journal of the American Academy of Child & Adolescent Psychiatry, 49(3), 229–238. 10.1016/j.jaac.2009.11.008

Biederman, J., Faraone, S. V., Keenan, K., Knee, D., & Tsuang, M. T. (1990). Family-genetic and psychosocial risk factors in DSM-III attention deficit disorder. Journal of the American Academy of Child & Adolescent Psychiatry, 29(4), 526–533.

Biederman, J., Petty, C. R., O’Connor, K. B., Hyder, L. L., & Faraone, S. V. (2012). Predictors of persistence in girls with attention deficit hyperactivity disorder: results from an 11-year controlled follow-up study. Acta Psychiatrica Scandinavica, 125(2), 147–156. 10.1111/j.1600-0447.2011.01797.x

Breiman, L. (2001). Random Forests. Machine Learning, 45(1), 5–32. 10.1023/A:1010933404324

Brown, M. R., Sidhu, G. S., Greiner, R., Asgarian, N., Bastani, M., Silverstone, P. H., Greenshaw, A. J., & Dursun, S. M. (2012). ADHD-200 Global Competition: diagnosing ADHD using personal characteristic data can outperform resting state fMRI measurements [Original Research]. Frontiers in Systems Neuroscience, Volume 6–2012. 10.3389/fnsys.2012.00069

Cao, M., Martin, E., & Li, X. (2023). Machine learning in attention-deficit/hyperactivity disorder: new approaches toward understanding the neural mechanisms. Translational Psychiatry, 13(1), 236. 10.1038/s41398-023-02536-w

Casey, B. J., Cannonier, T., Conley, M. I., Cohen, A. O., Barch, D. M., Heitzeg, M. M., Soules, M. E., Teslovich, T., Dellarco, D. V., Garavan, H., Orr, C. A., Wager, T. D., Banich, M. T., Speer, N. K., Sutherland, M. T., Riedel, M. C., Dick, A. S., Bjork, J. M., Thomas, K. M., . . . Dale, A. M. (2018). The Adolescent Brain Cognitive Development (ABCD) study: Imaging acquisition across 21 sites. Dev Cogn Neurosci, 32, 43–54. 10.1016/j.dcn.2018.03.001

Castellanos, F. X., Sharp, W. S., Gottesman, R. F., Greenstein, D. K., Giedd, J. N., & Rapoport, J. L. (2003). Anatomic Brain Abnormalities in Monozygotic Twins Discordant for Attention Deficit Hyperactivity Disorder. American Journal of Psychiatry, 160(9), 1693–1696. 10.1176/appi.ajp.160.9.1693

Chaim-Avancini, T. M., Doshi, J., Zanetti, M. V., Erus, G., Silva, M. A., Duran, F. L. S., Cavallet, M., Serpa, M. H., Caetano, S. C., Louza, M. R., Davatzikos, C., & Busatto, G. F. (2017). Neurobiological support to the diagnosis of ADHD in stimulant-naïve adults: pattern recognition analyses of MRI data. Acta Psychiatrica Scandinavica, 136(6), 623–636. 10.1111/acps.12824

Chen, Y. C., Sudre, G., Sharp, W., Donovan, F., Chandrasekharappa, S. C., Hansen, N., Elnitski, L., & Shaw, P. (2018). Neuroanatomic, epigenetic and genetic differences in monozygotic twins discordant for attention deficit hyperactivity disorder. Mol Psychiatry, 23(3), 683–690. 10.1038/mp.2017.45

Cheng, W., Ji, X., Zhang, J., & Feng, J. (2012). Individual classification of ADHD patients by integrating multiscale neuroimaging markers and advanced pattern recognition techniques [Article]. Frontiers in Systems Neuroscience(AUG 2012), Article 58. 10.3389/fnsys.2012.00058

Chiang, H.-L., Chen, Y.-J., Shang, C.-Y., Tseng, W.-Y., & Gau, S.-F. (2016). Different neural substrates for executive functions in youths with ADHD: a diffusion spectrum imaging tractography study. Psychological medicine, 46(6), 1225–1238.

Chiang, H.-L., Tseng, W.-Y. I., Tseng, W.-L., Tung, Y.-H., Hsu, Y.-C., Chen, C.-L., & Gau, S. S.-F. (2023). Atypical development in white matter microstructures in ADHD: A longitudinal diffusion imaging study. Asian Journal of Psychiatry, 79, 103358. 10.1016/j.ajp.2022.103358

Chiang, H. L., Tseng, W. I., Tseng, W. L., Tung, Y. H., Hsu, Y. C., Chen, C. L., & Gau, S. S. (2023). Atypical development in white matter microstructures in ADHD: A longitudinal diffusion imaging study. Asian J Psychiatr, 79, 103358. 10.1016/j.ajp.2022.103358

Colby, J. B., Rudie, J. D., Brown, J. A., Douglas, P. K., Cohen, M. S., & Shehzad, Z. (2012). Insights into multimodal imaging classification of ADHD. Front Syst Neurosci, 6, 59. 10.3389/fnsys.2012.00059

Dey, S., Rao, A. R., & Shah, M. (2012). Exploiting the brain’s network structure in identifying ADHD subjects. Front Syst Neurosci, 6, 75. 10.3389/fnsys.2012.00075

Fair, D. A., Nigg, J. T., Iyer, S., Bathula, D., Mills, K. L., Dosenbach, N. U., Schlaggar, B. L., Mennes, M., Gutman, D., & Bangaru, S. (2013). Distinct neural signatures detected for ADHD subtypes after controlling for micro-movements in resting state functional connectivity MRI data. Frontiers in Systems Neuroscience, 6, 80.

Fair, D. A., Posner, J., Nagel, B. J., Bathula, D., Dias, T. G., Mills, K. L., Blythe, M. S., Giwa, A., Schmitt, C. F., & Nigg, J. T. (2010). Atypical default network connectivity in youth with attention-deficit/hyperactivity disorder. Biol Psychiatry, 68(12), 1084–1091. 10.1016/j.biopsych.2010.07.003

Faraone, S. V., Banaschewski, T., Coghill, D., Zheng, Y., Biederman, J., Bellgrove, M. A., Newcorn, J. H., Gignac, M., Al Saud, N. M., Manor, I., Rohde, L. A., Yang, L., Cortese, S., Almagor, D., Stein, M. A., Albatti, T. H., Aljoudi, H. F., Alqahtani, M. M. J., Asherson, P., . . . Wang, Y. (2021). The World Federation of ADHD International Consensus Statement: 208 Evidence-based conclusions about the disorder. Neurosci Biobehav Rev, 128, 789–818. 10.1016/j.neubiorev.2021.01.022

Fischl, B. (2012). FreeSurfer. Neuroimage, 62(2), 774–781. 10.1016/j.neuroimage.2012.01.021

Fisher, A., Rudin, C., & Dominici, F. (2019). All Models are Wrong, but Many are Useful: Learning a Variable’s Importance by Studying an Entire Class of Prediction Models Simultaneously. J Mach Learn Res, 20.

Gau, S., Tseng, W.-L., Tseng, W.-Y., Wu, Y.-H., & Lo, Y.-C. (2015). Association between microstructural integrity of frontostriatal tracts and school functioning: ADHD symptoms and executive function as mediators. Psychological medicine, 45(3), 529–543.

González-Madruga, K., Staginnus, M., & Fairchild, G. (2022). Alterations in Structural and Functional Connectivity in ADHD: Implications for Theories of ADHD. In S. C. Stanford & E. Sciberras (Eds.), New Discoveries in the Behavioral Neuroscience of Attention-Deficit Hyperactivity Disorder (pp. 445–481). Springer International Publishing. 10.1007/7854_2022_345

Guyon, I., & Elisseeff, A. (2003). An introduction to variable and feature selection. J. Mach. Learn. Res., 3(null), 1157–1182.

Hagler Jr, D. J., Ahmadi, M. E., Kuperman, J., Holland, D., McDonald, C. R., Halgren, E., & Dale, A. M. (2009). Automated white-matter tractography using a probabilistic diffusion tensor atlas: Application to temporal lobe epilepsy. Human brain mapping, 30(5), 1535–1547.

He, N., Li, F., Li, Y., Guo, L., Chen, L., Huang, X., Lui, S., & Gong, Q. (2015). Neuroanatomical deficits correlate with executive dysfunction in boys with attention deficit hyperactivity disorder. Neurosci Lett, 600, 45–49. 10.1016/j.neulet.2015.05.062

Hoogman, M., Bralten, J., Hibar, D. P., Mennes, M., Zwiers, M. P., Schweren, L. S., van Hulzen, K. J., Medland, S. E., Shumskaya, E., & Jahanshad, N. (2017). Subcortical brain volume differences in participants with attention deficit hyperactivity disorder in children and adults: a cross-sectional mega-analysis. The Lancet Psychiatry, 4(4), 310–319.

Iannaccone, R., Hauser, T. U., Ball, J., Brandeis, D., Walitza, S., & Brem, S. (2015). Classifying adolescent attention-deficit/hyperactivity disorder (ADHD) based on functional and structural imaging. European Child & Adolescent Psychiatry, 24(10), 1279–1289. 10.1007/s00787-015-0678-4

Johnston, B. A., Mwangi, B., Matthews, K., Coghill, D., Konrad, K., & Steele, J. D. (2014). Brainstem abnormalities in attention deficit hyperactivity disorder support high accuracy individual diagnostic classification. Human Brain Mapping, 35(10), 5179–5189. 10.1002/hbm.22542

Kingma, D. P., & Ba, J. (2014). Adam: A method for stochastic optimization. *arXiv preprint arXiv:1412*.6980.

Konrad, K., Neufang, S., Hanisch, C., Fink, G. R., & Herpertz-Dahlmann, B. (2006). Dysfunctional attentional networks in children with attention deficit/hyperactivity disorder: evidence from an event-related functional magnetic resonance imaging study. Biological psychiatry, 59(7), 643–651.

Kumar, U., Arya, A., & Agarwal, V. (2021). Neural network connectivity in ADHD children: an independent component and functional connectivity analysis of resting state fMRI data. Brain Imaging Behav, 15(1), 157–165. 10.1007/s11682-019-00242-0

Larsson, H., Asherson, P., Chang, Z., Ljung, T., Friedrichs, B., Larsson, J.-O., & Lichtenstein, P. (2013). Genetic and environmental influences on adult attention deficit hyperactivity disorder symptoms: a large Swedish population-based study of twins. Psychological medicine, 43(1), 197–207.

LeCun, Y., Bengio, Y., & Hinton, G. (2015). Deep learning. nature, 521(7553), 436–444. 10.1038/nature14539

Loh, H. W., Ooi, C. P., Oh, S. L., Barua, P. D., Tan, Y. R., Molinari, F., March, S., Acharya, U. R., & Fung, D. S. S. (2023). Deep neural network technique for automated detection of ADHD and CD using ECG signal. Comput Methods Programs Biomed, 241, 107775. 10.1016/j.cmpb.2023.107775

Luo, Y., Alvarez, T. L., Halperin, J. M., & Li, X. (2020). Multimodal neuroimaging-based prediction of adult outcomes in childhood-onset ADHD using ensemble learning techniques. Neuroimage Clin, 26, 102238. 10.1016/j.nicl.2020.102238

Luo, Y., Alvarez, T. L., Halperin, J. M., & Li, X. (2020). Multimodal neuroimaging-based prediction of adult outcomes in childhood-onset ADHD using ensemble learning techniques. NeuroImage: Clinical, 26, 102238. 10.1016/j.nicl.2020.102238

Luo, Y., Halperin, J. M., & Li, X. (2020). Anatomical substrates of symptom remission and persistence in young adults with childhood attention deficit/hyperactivity disorder. European Neuropsychopharmacology, 33, 117–125. 10.1016/j.euroneuro.2020.01.014

Luo, Y., Weibman, D., Halperin, J. M., & Li, X. (2019). A Review of Heterogeneity in Attention Deficit/Hyperactivity Disorder (ADHD) [Review]. Frontiers in Human Neuroscience, 13. 10.3389/fnhum.2019.00042

Mahone, E. M., Ranta, M. E., Crocetti, D., O’Brien, J., Kaufmann, W. E., Denckla, M. B., & Mostofsky, S. H. (2011). Comprehensive examination of frontal regions in boys and girls with attention-deficit/hyperactivity disorder. Journal of the International Neuropsychological Society, 17(6), 1047–1057.

Marcus, D. S., Harms, M. P., Snyder, A. Z., Jenkinson, M., Wilson, J. A., Glasser, M. F., Barch, D. M., Archie, K. A., Burgess, G. C., Ramaratnam, M., Hodge, M., Horton, W., Herrick, R., Olsen, T., McKay, M., House, M., Hileman, M., Reid, E., Harwell, J., . . . Van Essen, D. C. (2013). Human Connectome Project informatics: quality control, database services, and data visualization. Neuroimage, 80, 202–219. 10.1016/j.neuroimage.2013.05.077

Martine Hoogman, Ph.D., Ryan Muetzel, Ph.D., Joao P. Guimaraes, M.Sc., Elena Shumskaya, Ph.D., Maarten Mennes, Ph.D., Marcel P. Zwiers, Ph.D., Neda Jahanshad, Ph.D., Gustavo Sudre, Ph.D., Thomas Wolfers, Ph.D., Eric A. Earl, B.Sc., Juan Carlos Soliva Vila, Ph.D., Yolanda Vives-Gilabert, Ph.D., Sabin Khadka, M.Sc., Stephanie E. Novotny, M.Sc., Catharina A. Hartman, Ph.D., Dirk J. Heslenfeld, Ph.D., Lizanne J.S. Schweren, Ph.D., Sara Ambrosino, M.D., Bob Oranje, Ph.D., . . . Barbara Franke, Ph.D. (2019). Brain Imaging of the Cortex in ADHD: A Coordinated Analysis of Large-Scale Clinical and Population-Based Samples. American Journal of psychiatry, 176(7), 531–542. 10.1176/appi.ajp.2019.18091033

Moon, M., & Nakai, K. (2016). Stable feature selection based on the ensemble L1-norm support vector machine for biomarker discovery. BMC Genomics, 17(13), 1026. 10.1186/s12864-016-3320-z

Mu, S., Wu, H., Zhang, J., & Chang, C. (2023). Subcortical structural covariance predicts symptoms in children with different subtypes of ADHD. Cerebral Cortex, 33(14), 8849–8857. 10.1093/cercor/bhad165

Nair, V., & Hinton, G. E. (2010). *Rectified linear units improve restricted boltzmann machines* Proceedings of the 27th International Conference on International Conference on Machine Learning, Haifa, Israel.

Peterson, D. J., Ryan, M., Rimrodt, S. L., Cutting, L. E., Denckla, M. B., Kaufmann, W. E., & Mahone, E. M. (2011). Increased regional fractional anisotropy in highly screened attention-deficit hyperactivity disorder (ADHD). J Child Neurol, 26(10), 1296–1302. 10.1177/0883073811405662

Polanczyk, G. V., Willcutt, E. G., Salum, G. A., Kieling, C., & Rohde, L. A. (2014). ADHD prevalence estimates across three decades: an updated systematic review and meta-regression analysis. International Journal of Epidemiology, 43(2), 434–442. 10.1093/ije/dyt261

Qiu, M.-g., Ye, Z., Li, Q.-y., Liu, G.-j., Xie, B., & Wang, J. (2011). Changes of brain structure and function in ADHD children. Brain topography, 24(3), 243–252.

Qureshi, M. N. I., Min, B., Jo, H. J., & Lee, B. (2016). Multiclass classification for the differential diagnosis on the ADHD subtypes using recursive feature elimination and hierarchical extreme learning machine: structural MRI study. PLOS ONE, 11(8), e0160697.

Qureshi, M. N. I., Oh, J., Min, B., Jo, H. J., & Lee, B. (2017). Multi-modal, multi-measure, and multi-class discrimination of ADHD with hierarchical feature extraction and extreme learning machine using structural and functional brain MRI. Frontiers in human neuroscience, 11, 157.

Saad, J. F., Griffiths, K. R., & Korgaonkar, M. S. (2020). A Systematic Review of Imaging Studies in the Combined and Inattentive Subtypes of Attention Deficit Hyperactivity Disorder [Systematic Review]. Frontiers in Integrative Neuroscience, 14. 10.3389/fnint.2020.00031

Samuel, V. J., George, P., Thornell, A., Curtis, S., Taylor, A., Brome, D., Mick, E., Faraone, S. V., & Biederman, J. (1999). A pilot controlled family study of DSM-III-R and DSM-IV ADHD in African-American children. Journal of the American Academy of Child & Adolescent Psychiatry, 38(1), 34–39.

Sen, B., Borle, N. C., Greiner, R., & Brown, M. R. G. (2018). A general prediction model for the detection of ADHD and Autism using structural and functional MRI. PLOS ONE, 13(4), e0194856. 10.1371/journal.pone.0194856

Sprich, S., Biederman, J., Crawford, M. H., Mundy, E., & Faraone, S. V. (2000). Adoptive and biological families of children and adolescents with ADHD. Journal of the American Academy of Child & Adolescent Psychiatry, 39(11), 1432–1437.

Strobl, C., Boulesteix, A.-L., Kneib, T., Augustin, T., & Zeileis, A. (2008). Conditional variable importance for random forests. BMC Bioinformatics, 9(1), 307. 10.1186/1471-2105-9-307

Sun, Y., Zhao, L., Lan, Z., Jia, X. Z., & Xue, S. W. (2020). Differentiating Boys with ADHD from Those with Typical Development Based on Whole-Brain Functional Connections Using a Machine Learning Approach. Neuropsychiatr Dis Treat, 16, 691–702. 10.2147/ndt.S239013

Tang, Y., Wang, C., Chen, Y., Sun, N., Jiang, A., & Wang, Z. (2021). Identifying ADHD Individuals From Resting-State Functional Connectivity Using Subspace Clustering and Binary Hypothesis Testing. Journal of Attention Disorders, 25(5), 736–748. 10.1177/1087054719837749

Tung, Y.-H., Lin, H.-Y., Chen, C.-L., Shang, C.-Y., Yang, L.-Y., Hsu, Y.-C., Tseng, W.-Y. I., & Gau, S. S.-F. (2021). Whole brain white matter tract deviation and idiosyncrasy from normative development in autism and ADHD and unaffected siblings link with dimensions of psychopathology and cognition. American Journal of Psychiatry, 178(8), 730–743.

van’t Ent, D., van Beijsterveldt, C. E., Derks, E. M., Hudziak, J. J., Veltman, D. J., Todd, R. D., Boomsma, D. I., & De Geus, E. J. (2009). Neuroimaging of response interference in twins concordant or discordant for inattention and hyperactivity symptoms. Neuroscience, 164(1), 16–29. 10.1016/j.neuroscience.2009.01.056

Vergara, J. R., & Estévez, P. A. (2014). A review of feature selection methods based on mutual information. Neural Computing and Applications, 24(1), 175–186. 10.1007/s00521-013-1368-0

Xia, S., Li, X., Kimball, A. E., Kelly, M. S., Lesser, I., & Branch, C. (2012). Thalamic shape and connectivity abnormalities in children with attention-deficit/hyperactivity disorder. Psychiatry Research: Neuroimaging, 204(2), 161–167. 10.1016/j.pscychresns.2012.04.011

Yasumura, A., Omori, M., Fukuda, A., Takahashi, J., Yasumura, Y., Nakagawa, E., Koike, T., Yamashita, Y., Miyajima, T., & Koeda, T. (2020). Applied machine learning method to predict children with ADHD using prefrontal cortex activity: a multicenter study in Japan. Journal of Attention Disorders, 24(14), 2012–2020.

Yu, M., Gao, X., Niu, X., Zhang, M., Yang, Z., Han, S., Cheng, J., & Zhang, Y. (2022). Meta-analysis of structural and functional alterations of brain in patients with attention-deficit/hyperactivity disorder. Front Psychiatry, 13, 1070142. 10.3389/fpsyt.2022.1070142

Zöllei, L., Iglesias, J. E., Ou, Y., Grant, P. E., & Fischl, B. (2020). Infant FreeSurfer: An automated segmentation and surface extraction pipeline for T1-weighted neuroimaging data of infants 0–2 years. Neuroimage, 218, 116946. 10.1016/j.neuroimage.2020.116946

Zou, L., Zheng, J., Miao, C., Mckeown, M. J., & Wang, Z. J. (2017). 3D CNN based automatic diagnosis of attention deficit hyperactivity disorder using functional and structural MRI. Ieee Access, 5, 23626–23636.

